# AngioNet: A Convolutional Neural Network for Vessel Segmentation in X-ray Angiography

**DOI:** 10.1101/2021.01.25.21250488

**Authors:** Kritika Iyer, Cyrus P. Najarian, Aya A. Fattah, Christopher J. Arthurs, S.M. Reza Soroushmehr, Vijayakumar Subban, Mullasari A. Sankardas, Raj R. Nadakuditi, Brahmajee K. Nallamothu, C. Alberto Figueroa

## Abstract

Coronary Artery Disease (CAD) is commonly diagnosed using X-ray angiography, in which images are taken as radio-opaque dye is flushed through the coronary vessels to visualize stenosis severity. Cardiologists typically use visual estimation to approximate the percent diameter reduction of the stenosis, and this directs therapies like stent placement. A fully automatic method to segment the vessels would eliminate potential subjectivity and provide a quantitative and systematic measurement of diameter reduction. Here, we have designed a convolutional neural network, AngioNet, for vessel segmentation in X-ray angiography images. The main innovation in this network is the introduction of an Angiographic Processing Network which significantly improves segmentation performance on multiple network backbones, with the best performance using Deeplabv3+ (Dice score 0.864, sensitivity 0.918, specificity 0.987). We have also demonstrated the interchangeability of our network in measuring vessel diameter with Quantitative Coronary Angiography. Our results indicate that AngioNet is a powerful tool for automatic angiographic vessel segmentation that could facilitate systematic anatomical assessment of coronary stenosis in the clinical workflow.

## Introduction

Coronary Artery Disease (CAD) affects over 20 million adults in the United States and accounts for nearly one-third of adult deaths in western countries ^1–3^. The annual cost to the United States healthcare system for the first year of treatment is $5.54 billion ^4^. The disease is characterized by the buildup of plaque in the coronary arteries ^5,6^, which causes a narrowing of the blood vessel known as stenosis.

CAD is most commonly diagnosed using X-ray angiography ^7^, whereby a catheter is inserted into the patient and a sequence of X-ray images are taken as radio-opaque dye is flushed into the coronary arteries. Cardiologists typically approximate stenosis severity via visual inspection of the X-ray images, estimating the percent reduction in diameter or cross-sectional area. If the area reduction is believed to be greater than 70%, a revascularization procedure, such as stent placement or coronary artery bypass grafting surgery, may be performed to treat the stenosis ^8,9^.

Quantitative Coronary Angiography, or QCA, is a diagnostic tool used in conjunction with X-ray angiography to more accurately determine stenosis severity ^10,11^. QCA is an accepted standard for assessment of coronary artery dimensions and uses semi-automatic edge-detection algorithms to quantify the change in vessel diameter. The QCA software then reports the diameter at user-specified locations as well as the percentage diameter reduction at the stenosis ^12^. Although QCA is more quantitative than visual inspection alone, it requires human input and time to identify the stenosis and to manually correct the vessel boundaries before calculating the stenosis percentage. This has led to QCA largely being used in the setting of clinical studies with limited impact on patient care. A fully automatic angiographic segmentation algorithm would speed up the process of determining stenosis severity, eliminate the need for subjective manual corrections, and potentially lead to broader utilization in clinical workflows.

Fully-automated angiographic segmentation is particularly challenging due to the poor signal-to-noise ratio and overlapping structures such as the catheter and the patient’s spine and rib cage ^13^. Several filter-based approaches^13–22^ and convolutional neural networks (CNNs) ^23–28^ have been developed for angiographic segmentation. While many of these approaches had success in isolating the coronary vessels, many are time-consuming and require manual correction, while others are limited in their ability to separate the vessels from other structures such as the catheter.

To address these shortcomings, we have developed a new CNN for angiographic segmentation: AngioNet, which combines an Angiographic Processing Network (APN) with a semantic segmentation network. The APN was trained to address several of the challenges specific to angiographic segmentation, including low contrast images, presence of the catheter, and overlapping bony structures. AngioNet uses Deeplabv3+ as its backbone semantic segmentation network instead of U-Net or other simpler fully convolutional networks (FCNs), which are more commonly used for medical segmentation. The deeper architecture of Deeplabv3+ compared to the FCNs typically used for medical segmentation allows our network to learn more features and perform well in challenging cases. In this paper, we also explored the specific benefits of the APN – and the importance of using Deeplabv3+ as the backbone – by comparing segmentation accuracy in Deeplabv3+ and U-Net, trained both with and without the APN. Lastly, we performed clinical validation of segmentation accuracy by comparing AngioNet-derived vessel diameter against QCA-derived diameter.

## Results

### Patient Characteristics

#### 1. UM Dataset

This dataset was composed of de-identified angiograms acquired using a Siemens Artis Q Angiography system at the University of Michigan (UM) Hospital. The enrollment criterion for this dataset was patients referred for a diagnostic coronary angiography procedure done at the UM Hospital in 2017. Patients with pacemakers, implantable defibrillators, or prior cardiac surgery with coronary artery bypass grafts were excluded, as these prior procedures introduce artifacts and additional vascular conduits. Furthermore, patients with diffuse stenosis were excluded as this is less common in arteries suitable for revascularization and is a challenging case for segmentation. In our sample of 161 patients, 14 had severe stenosis (≤ 80% diameter reduction) and the remaining had mild to moderate stenosis. The dataset was composed of 280 images of the left coronary artery (LCA) and 182 images of the right coronary artery (RCA), which were equally split into 6 partitions to generate a 5-fold cross validation dataset and a test set.

#### 2. MMM QCA Dataset

The Madras Medical Mission (MMM) QCA dataset contained independently generated three-vessel QCA reports of 89 patients, encompassing 223 vessels in both the LCA and RCA. All patients presented with mild to moderate stenosis. The data were acquired from the Indian Cardiovascular Core Laboratory (ICRF) at the MMM, which serves as a core laboratory with experience in clinical trials and other studies and has expertise in QCA.

### Learned Filters using the Angiographic Processing Network

The main contribution of our work is the development of an Angiographic Processing Network (APN). The purpose of the APN is to learn the best possible preprocessing filter that will improve segmentation performance, incorporating the characteristics of boundary sharpening and contrast enhancing filters. The APN works in tandem with segmentation neural networks (backbone networks) to create a composite neural network such as AngioNet, a combination of the APN and Deeplabv3+.

Figure 1 contains examples of the filters that the APN learned when it was trained with Deeplabv3+ or U-Net, respectively. The images represent the output of the APN, and thus the input to the backbone network. Although the APN was initialized with the combination of unsharp mask filters shown in Figure 1, the network learns different filters that perform a combination of contrast-enhancement and boundary sharpening. The examples given are the results of training with different data partitions during k-fold cross-validation. The large variations in the learned filters come from an inherent property of neural network training; since minimization of the neural network’s loss function is a non-convex optimization problem ^44^, there are many combinations of network weights which will lead to similar results. The effect of these varied learned filters on segmentation accuracy is described in the next section.

**Figure 1.**
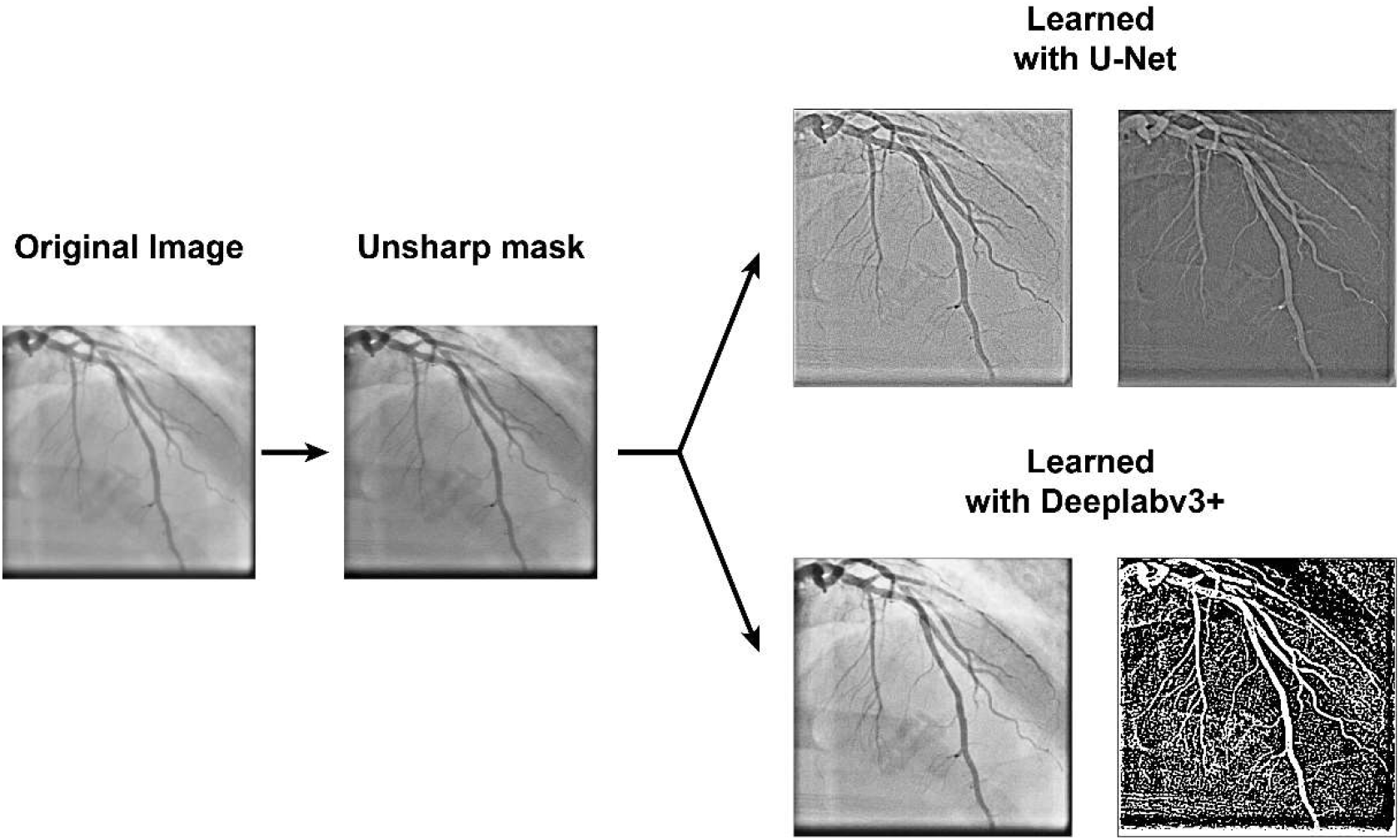
Examples of learned filters when the APN is trained with Deeplabv3+ and U-Net. The APN was initialized with the combination of unsharp mask filters shown above, and learned new filters to aid segmentation. Each example image is the output of the APN after training with different data partitions.

### Comparison of AngioNet versus Current State-of-the-art Semantic Segmentation Neural Networks

Segmentation accuracy was measured using the Dice score, given by

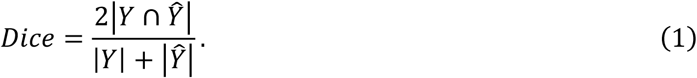

Here, *Y* is the label image and *Ŷ* is the neural network prediction, each of which is a binary image where vessel pixels have a value of 1 and background pixels have a value of 0. | *Y* | denotes the number of vessel pixels (1s) in image *Y*, and ∩ represents a pixel-wise logical AND operation. Alternatively, the Dice score can be defined in terms of the true positives (TP), false positives (FP) and false negatives (FN) of the neural network prediction with respect to the label image, and is then given by

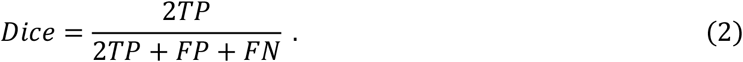

We also report the Area under the Receiver-Operator Curve (AUC), which measures the ability of the network to separate classes, in this case, vessel and background pixels. An AUC of 0.5 indicates a model that is no better than random chance, whereas an AUC of 1 indicates a model that can perfectly discriminate between classes. Finally, we also report the pixel accuracy of the binary segmentation, defined as

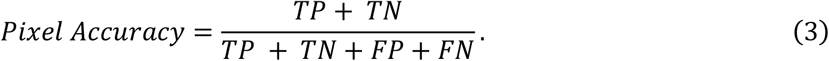

#### 1. UM Dataset

The accuracy of AngioNet was validated using a 5-fold cross-validation study, in which the neural network was trained on 4 out of the 5 training data partitions at a time, with the fifth partition reserved for validation and hyperparameter optimization (hold-out set). This process was repeated five times, holding out a different partition each time. The accuracy of the resulting five trained networks was measured on the sixth partition, the test set, which was never used for training. The mean k-fold accuracy and the accuracy when trained on all five training data partitions are summarized in Table 1. The network performs well on both LCA and RCA input images (Figure 2) and does not segment the catheter or other imaging artifacts despite uneven brightness, overlapping structures, and varying contrast.

**Table 1:** Accuracy of AngioNet using K-Fold Cross Validation

|  | **Dice Score** | **Sensitivity** | **Specificity** | **AUC** | **Pixel Accuracy** |
| --- | --- | --- | --- | --- | --- |
| **k-Fold Test** | 0.856±0.004 | 0.913±0.013 | 0.987±0.001 | 0.991±0.002 | 0.982±0.004 |
| **k-Fold Holdout** | 0.857±0.012 | 0.909±0.012 | 0.987±0.001 | 0.990±0.002 | 0.980±0.003 |
| **All Data** | 0.864 | 0.918 | 0.987 | 0.991 | 0.983 |

**Figure 2.**
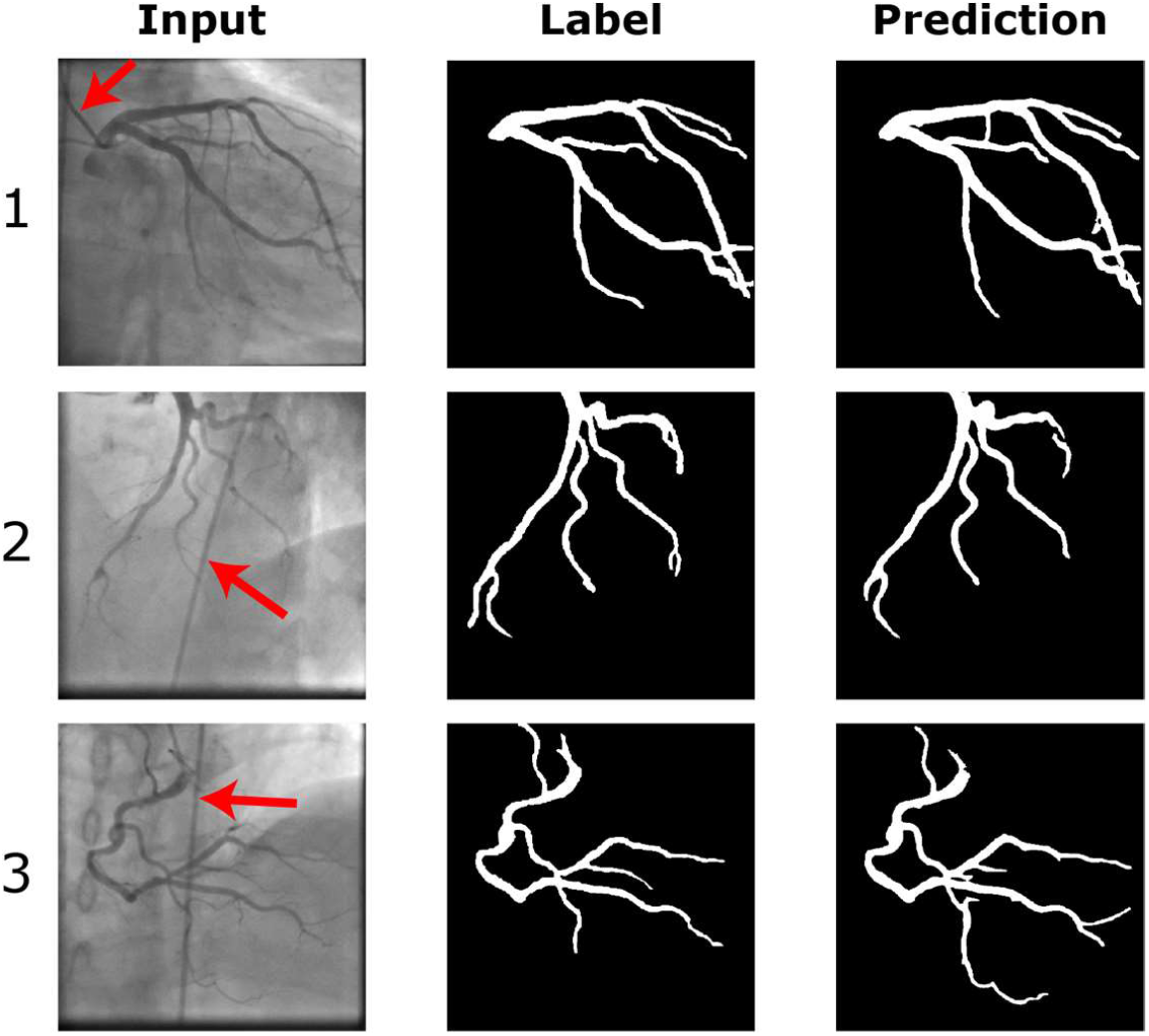
Examples of AngioNet segmentation on left coronary tree, taken at two different angles (1,2), and right coronary tree (3). AngioNet does not segment the catheter (red arrows), despite its similar diameter and pixel intensity as the vessels (2,3). It also ignores bony structures such as the spine in (3) and ribs in (1).

The Dice score distribution on the test set for AngioNet, Deeplabv3+, U-Net with the APN (APN+U-Net), and U-Net is shown in Figure 3A. All networks were trained using the UM Dataset. AngioNet has the highest mean Dice score on the test set (0.864) when trained on all five partitions of the training data, compared to 0.815 for Deeplabv3+ alone, 0.811 for APN+U-Net, and 0.717 for U-Net alone. On average, AngioNet has a 10% higher Dice score per image than Deeplabv3+ alone. APN+U-Net has a 14% higher Dice score than U-Net alone. A paired Student’s t-test was performed to determine if the addition of the APN significantly improved the Dice score for both network backbones. The p-value between AngioNet and Deeplabv3+ was 5.76e-10, while the p-value between APN+U-Net and U-Net was 2.63e-16. Both p-values were much less than the statistical significance threshold of 0.05, therefore we can conclude that there are statistically significant differences between the Dice score distributions with and without the APN. Furthermore, both Deeplabv3+ and U-Net exhibit outliers with Dice score lower than 0.4, but adding the APN eliminates these outliers in both networks.

**Figure 3.**
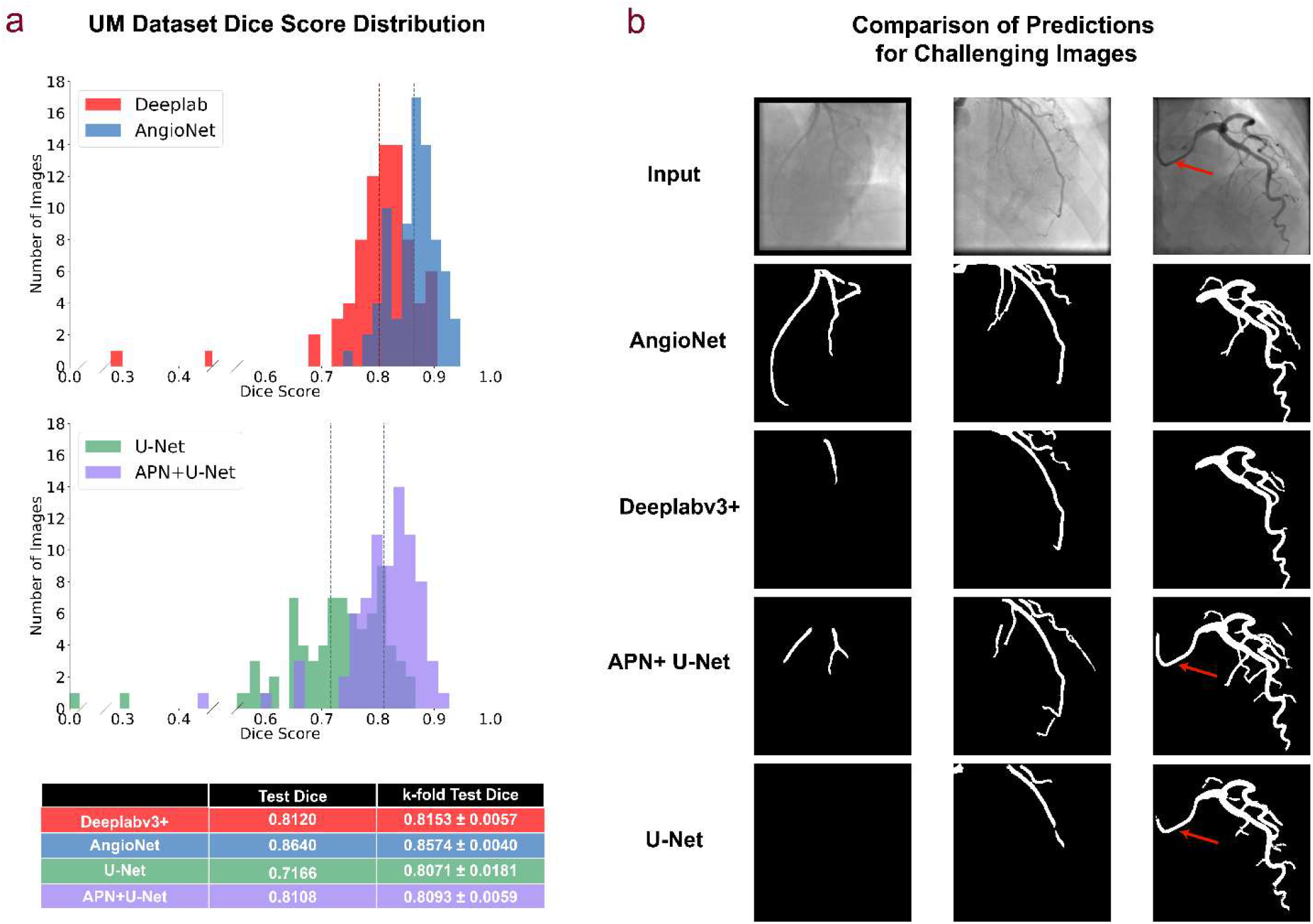
Summary of AngioNet (APN + Deeplabv3+) performance. All results are derived from the networks trained on all five partitions of the UM training set, unless otherwise noted as a k-fold result. a) Comparison of Dice score distribution on test set. AngioNet has the highest average Dice score, with scores ranging from 0.737 to 0.946. Adding the APN improves the lowest Dice scores of both Deeplabv3+ and U-Net. Dashed lines correspond to the Test Dice in the table below. b) Segmentation comparison on challenging images with low contrast, faint vessels, and a curved catheter. AngioNet can segment more vessels in these images without segmenting the catheter (red arrows).

Compared to the other three networks, AngioNet performs the best on the challenging cases shown in Figure 3B. The first column of Figure 3B shows segmentation performance on a low contrast angiography image. AngioNet can segment the major coronary vessels in this image, whereas Deeplabv3+ only segments one vessel and U-Net is unable to identify any vessels at all. APN+U-Net can segment more vessels than U-Net alone, indicating once again that the addition of the APN improves segmentation performance on these low contrast images. In the second column, both AngioNet and APN+U-Net can segment fainter and smaller diameter vessels than the other two networks. Finally, we observe that AngioNet and Deeplabv3+ did not segment the catheter in the third column, although it is of similar diameter and curvature as the coronary vessels. Conversely, both APN+U-Net and U-Net included the catheter in their segmentations. Overall, AngioNet segmented the catheter in 2.6% of the images, where the catheter curved across the image and overlapped with the vessel. In contrast, Deeplabv3+ segmented the catheter in 6.4% of images. Both networks performed better than U-Net and APN+U-Net, which segmented the catheter in 19.5% and 9.1% of images respectively.

### Evaluation of Vessel Diameter Accuracy versus QCA

Evaluation of vessel diameter accuracy was done using the MMM QCA dataset. Maximum and minimum vessel diameter were compared in 255 vessels including both the RCA and LCA. On average, the absolute error in vessel diameter between the AngioNet segmentation and QCA report was 0.272mm or 1.15 pixels.

The linear regression plot in Figure 4A shows that vessel diameter estimates of both methods are linearly proportional and tightly clustered around the line of best fit, *y* = 0.957*x* − 0.106, Pearson’s correlation coefficient, *r* = 0.9866. The standardized difference ^45^, also known as Cohen’s effect size ^46^, was used to determine the difference in means between the diameter distributions of AngioNet and QCA. The standardized difference can determine significant differences between two groups in clinical studies^45^. The standardized difference between the AngioNet and QCA diameter distributions is 0.215, suggesting small differences between the two method.

**Figure 4.**
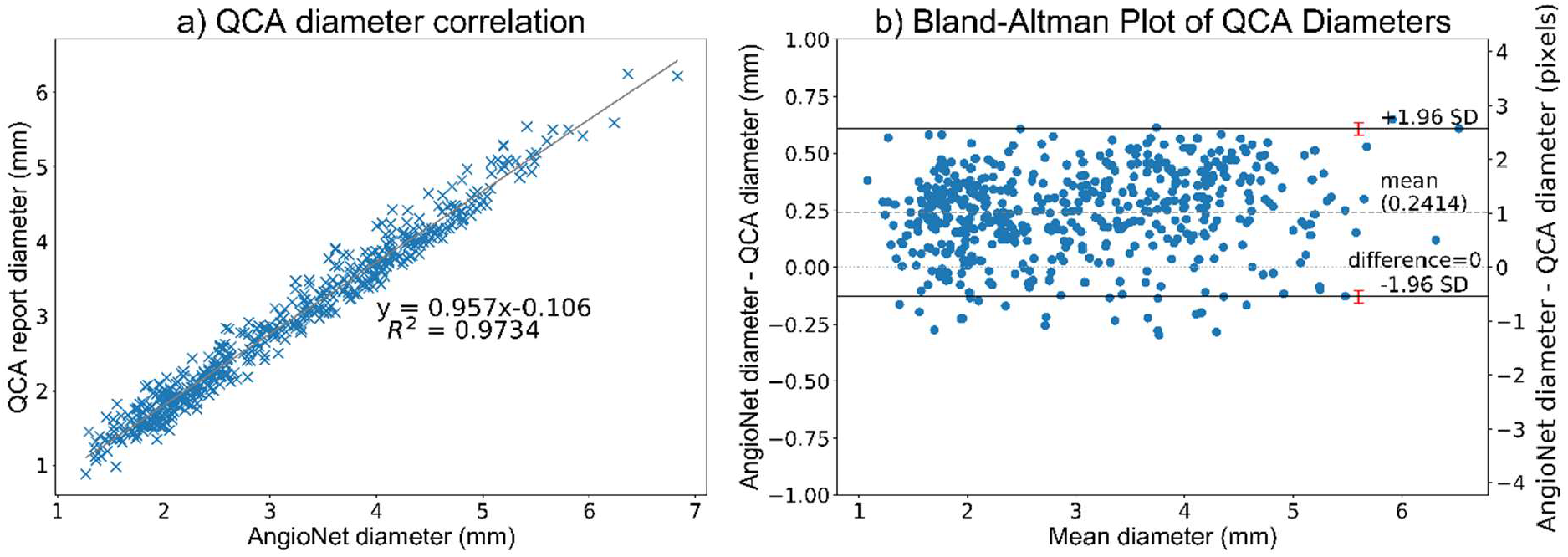
a) Correlation plot of QCA and AngioNet derived vessel diameters. b) The Bland-Altman plot demonstrates that AngioNet’s segmentation and QCA are interchangeable methods to determine vessel diameter since more than 95% of points lie within the limits of agreement. The red error bars represent the 95% confidence interval containing the limits of agreement. The mean difference in diameter between methods is 0.24mm or 1.1 pixels

Figure 4B is a Bland-Altman plot demonstrating the interchangeability of the QCA and AngioNet-derived diameters. The mean difference between both measures, 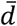, is 0.2414. The magnitude of the diameter difference remains relatively constant for all mean diameter values, indicating that there is a systematic error and not a proportional error between the two measurements. The limits of agreement are defined as 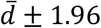, where SD is the standard deviation of the diameter differences. For both measurements to be considered interchangeable, 95% of data points must lie between these limits of agreement. In this plot, 96% of data points are within 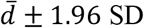. When including the 95% confidence interval of the limits of agreement as recommended by Bland and Altman ^47^, 97% of data points are within the range.

## Discussion

To put our work in context, the most common approaches to angiographic segmentation involve image processing techniques such as edge detection algorithms, matrix decomposition, wavelet-based methods, active contours, or the Frangi vesselness filter ^13–18^. Although these techniques can produce successful segmentations, they perform poorly on low-contrast or noisy images and cannot separate overlapping catheters and bony structures from the vessels. Several other approaches to angiographic segmentation such as Hessian-based random walk, region-growing, parametric density estimation, and deformation models have been explored ^19–22^; these semi-automatic methods still require manual correction or user input, ultimately hampering their applicability within the clinical workflow. To address these limitations, some have turned to convolutional neural networks (CNNs) for angiographic segmentation instead.

CNNs have been used for segmentation in numerous applications ^29–34^. Although originally designed for biomedical image segmentation, U-Net and its variations have been widely adopted in other fields due to their relatively simple architecture and high accuracy on binary segmentation problems ^36–38^. Despite its success, the network is not as deep as other state-of-the-art CNNs and thus lacks the expressive power to perform complex tasks such as angiographic segmentation ^39–41^. A more complex CNN for semantic segmentation is Deeplabv3+ ^42,43^. Although Deeplabv3+ performs well on multi-class segmentation tasks, it was not designed with the specific challenges of angiographic imaging in mind.

Several CNNs have been designed specifically for angiographic segmentation ^23–28^. Yang et al. ^23^ developed a CNN to segment the major branches of the coronary arteries. Despite its high segmentation accuracy, this network was only developed for single vessel segmentation. Multichannel Segmentation Network with Aligned input (MSN-A) ^24^, is a CNN based on U-Net designed to segment the entire coronary tree. The inputs to MSN-A are the angiographic image and a co-registered “mask” image taken before the dye was injected into the vessel. The main drawback of this network is that the multi-input strategy requires the entire angiographic sequence to be acquired with minimal table motion, whereas standard clinical practice involves moving the patient table to follow the flow of dye within the vessels. Nasr-Esfahani et al. ^25^ developed their own CNN architecture for angiographic segmentation, combining contrast enhancement, edge detection, and feature extraction. Shin et al. ^26^ combined a feature-extraction convolutional network with a graph convolutional network and inference CNN to create Vessel Graph Network (VGN) and improve segmentation performance by learning the tree structure of the vessels. Despite successfully segmenting most of the branches of the coronary tree, both of these methods had lower accuracy than MSN-A.

In the following sections, we will demonstrate that AngioNet is comparable to these state-of-the-art methods for angiographic segmentation. The key findings and clinical implications of our study are also described below.

### Learned Filters using the Angiographic Processing Network

As seen in Figure 1, the APN learns many different preprocessing filters that improve segmentation performance based on the data partition used for training. All learned filters exhibit both boundary sharpening and local contrast enhancement, likely due to the network’s initialization as a combination of unsharp mask filters. Despite the variation in the learned filters, the overall segmentation accuracy remains relatively constant as indicated by the small standard deviation from k-fold cross validation (0.004 for AngioNet, and 0.006 for APN+U-Net). This demonstrates that despite the large variation to the human eye, the different combinations of learned weights all achieved similar local minima of the loss function, leading to similar Dice scores. When compared to Deeplabv3+’s predictions on images preprocessed using unsharp masking filters, AngioNet’s segmentation accuracy is superior (0.864 for AngioNet, compared to 0.833 for Deeplabv3+). This suggests that the learned preprocessing filter implemented in this work is superior to manually selecting a particular contrast enhancement or boundary sharpening filter for preprocessing.

### Comparison of AngioNet to Current State-of-the-Art Semantic Segmentation Neural Networks

Several aspects of AngioNet’s design contribute to its enhanced segmentation performance compared to existing state of the art networks. The APN successfully improves segmentation performance on low contrast images compared to previous state-of-the-art semantic segmentation networks (Figure 3A). The APN also enhances performance on smaller vessels, which have lower contrast than larger vessels because they contain less radio-opaque dye. Without the APN, Deeplabv3+ and U-Net are not equipped to identify these faint vessels and underpredict the presence of small coronary branches. As seen in Figure 3A and Figure 3B, both Deeplabv3+ and AngioNet perform better than U-Net on angiographic segmentation. The addition of the APN to U-Net significantly increases the mean Dice score, facilitates segmentation of more vessels compared to U-Net alone, and greatly reduces the proportion of segmentation that include the catheter; yet APN+U-Net has some of the same drawbacks of U-Net such as disconnected vessels and more instances of the catheter being segmented compared to Deeplabv3+ and AngioNet. Although U-Net has demonstrated great success in other binary segmentation applications ^35,37^, the presence of catheters and bony structures with similar dimensions and pixel intensity as the vessels of interest make this a particularly challenging segmentation task. Deeplabv3+ and AngioNet have a deeper, more complex architecture, which allows these networks to learn more features with which to identify the vessels in each image ^39–41^.

The effective receptive field size U-Net is 64×64 pixels whereas that of Deeplabv3+ is 128×128 pixels ^48^. A larger receptive field is associated with better pixel localization and segmentation accuracy, as well as classification of larger scale objects in an image ^49,50^. Deeplabv3+’s larger receptive field may explain why Deeplabv3+ and AngioNet are more successful in avoiding segmentation of the catheter, an object typically larger than U-Net’s 64×64 pixel receptive field. The larger receptive field may also explain why Deeplabv3+ and AngioNet are better able to preserve the continuity of the coronary vessel tree and produce fewer broken or disconnected vessels than U-Net and APN+U-Net. Thus, Deeplabv3+ was an appropriate choice of network backbone for AngioNet.

AngioNet also demonstrates advantages compared to networks trained specifically for angiographic segmentation. Shin *et al*. reported a maximum Dice score of 0.82 and 0.837 on two datasets of retinal angiograms using VGN, whereas AngioNet’s maximum Dice score is 0.946 ^26^. Fan *et al*. reported Dice scores of 0.872 on a test set of 18 angiograms using MSN-A ^24^. AngioNet’s mean Dice score of 0.864 is very close to MSN-A’s 0.872. Although our mean Dice score is slightly lower than that of MSN-A, the major advantage of our network is that its input angiograms are not limited to those acquired with minimal movement of the patient table. AngioNet’s additional strengths compared to previous networks include ignoring overlapping structures when segmenting the coronary vessels, smaller sensitivity to noise, and the ability to segment low contrast images. The ability to avoid overlapping bony structures or the catheter is especially important as this eliminates the need for manual correction of the vessel boundary, which is a major advantage over mechanistic segmentation approaches.

AngioNet’s greatest limitation is that it overpredicts the vessel boundary in cases of severe (>85%) stenosis. The network performs well on mild and moderate stenoses, but it has learned to smooth the vessel boundary when the diameter sharply decreases to a single pixel. This is likely due to the low number of training examples containing severe stenosis: only 14 out of the 462 images in the entire UM Dataset contained severe stenosis, and two of these were in the test set. This drawback can be addressed by increasing the training data to encompass more examples of severe stenosis.

### Evaluation of Vessel Diameter Accuracy

A significant clinical implication of our findings was the comparison between AngioNet and QCA. In Figure 4A, we observe that QCA and AngioNet results are clustered around the line of best fit, *y* = 0.957*x* − 0.106. Given that the slope of the line of best fit is nearly 1, the intercept is close to 0, and the Pearson’s coefficient *r* is 0.9866, the line of best fit indicates strong agreement between these two methods of determining vessel diameter. The *R*^2^ coefficient for the linear regression model implies that 97.34% of the variance in the data can be explained by the line of best fit.

The standardized difference, or effect size, is a measure of how many pooled standard deviations separate the means of two distributions ^45^. According to Cohen, an effect size of 0.2 is considered a small difference between both groups, 0.5 is a medium difference, and 0.8 is a large difference ^46^. Given that the effect size between the QCA and AngioNet diameter distributions was 0.215 (91.5% overlap between the two distributions), we can conclude that the difference between QCA and AngioNet diameters are small. Furthermore, since the standardized difference indicated no large difference between QCA and AngioNet diameter estimations, these results suggest that both methods can be used interchangeably from a clinical perspective for the dataset examined.

The Bland-Altman plot in Figure 4B shows that the mean difference between QCA and AngioNet diameters is approximately 1.1 pixels. AngioNet under-predicts the vessel boundary by no more than 1 pixel and over-predicts by no more than 2.5 pixels. To put these values in context, the inter-operator variability for annotating the vessel boundary is 0.18±0.24mm or slightly above 1 pixel according to a study by Hernandez-Vela et al ^51^. The 95% confidence intervals of the limits of agreement were taken into consideration when determining how many data points lie between the limits of agreement as recommended by Bland and Altman ^47^. 97% of the data points lie within the range, which is greater than the 95% threshold. Given these results, and that the standardized difference test which produced no significant difference between the methods, one can conclude that QCA and AngioNet are interchangeable methods to determine vessel diameter. Given AngioNet’s fully automated nature, the workload required for generating QCA due to human input could be substantially reduced. Although our direct comparison of AngioNet-derived diameters with QCA-derived diameters required user interaction, future work will focus on developing an automated algorithm for stenosis detection and measurement based on the outputs of AngioNet’s segmentation.

In conclusion, AngioNet was designed to address the shortcomings of current state-of-the-art neural networks for X-ray angiographic segmentation. The APN was found to be a critical component to improve detection and segmentation of the coronary vessels, leading to 14% and 10% improved Dice score compared to U-Net or Deeplabv3+ alone. AngioNet demonstrated better segmentation accuracy than U-Net and Deeplabv3+, particularly on images with poor contrast or many small vessels. It also demonstrated increased robustness to ignoring the catheter and other imaging artifacts compared to other networks. Furthermore, our statistical analysis of the vessel diameters determined by AngioNet and traditional QCA demonstrated that the two methods may be interchangeable which could have large implications for clinical workflows. Future work to improve performance will focus on increasing accuracy on severe stenosis cases and automating stenosis measurement.

## Methods

### Datasets

Figure 5 summarizes the two patient datasets used in this work for neural network training and evaluation of performance against clinically relevant metrics and other state-of-the art networks. All data were collected in compliance with ethical guidelines.

**Figure 5.**
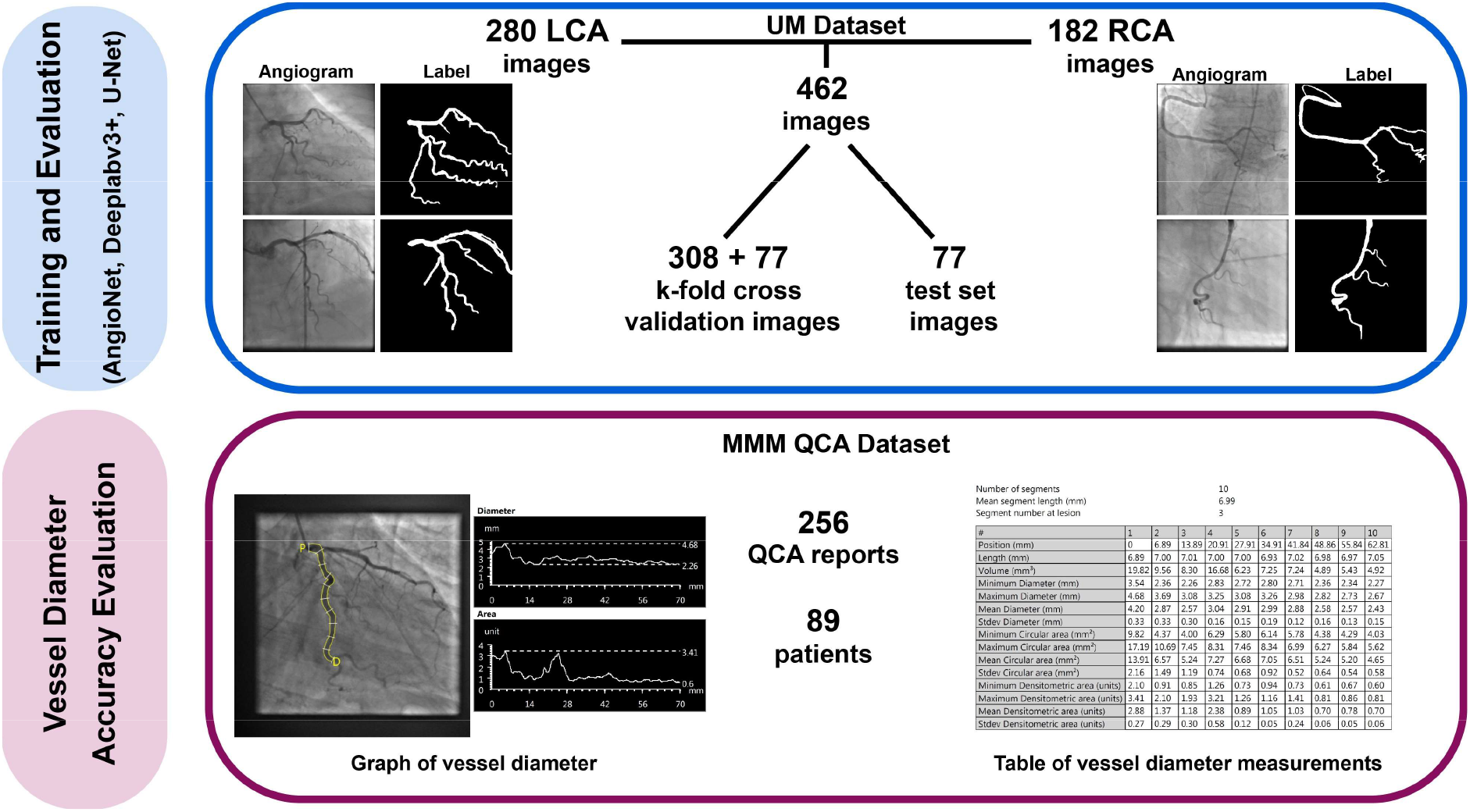
Diagram of datasets for CNN training and evaluation. AngioNet’s performance was compared against state-of-the-art neural networks, all trained on the UM Dataset. The MMM QCA dataset was used to quantify segmentation diameter accuracy by comparing AngioNet’s results against the diameters reported in QCA. Acronyms: left coronary artery (LCA); right coronary artery (RCA); Madras Medical Mission (MMM).

#### 1. UM Dataset

The UM Dataset was composed of 462 de-identified coronary angiograms acquired at the UM Hospital. The study protocol to access this data (HUM00084689) was reviewed by the Institutional Review Boards of the University of Michigan Medical School (IRBMED). Since the data was collected retrospectively, IRBMED approved use without requiring informed consent.

The data were equally split by patient into a 5-fold cross-validation set and test set to avoid having images from the same patient in both the training and test sets. Labels for all images were manually generated using Adobe Photoshop to include vessels with a diameter greater than 1mm at their origin. The 5-fold cross-validation portion of the dataset was used for neural network training and hyperparameter optimization, whereas the test set was used to evaluate segmentation accuracy.

There is a great number of artifacts in X-ray angiography images, including borders from X-ray filters, rotation of the image frame, varying levels of contrast, and magnification during image acquisition. Data augmentation of the UM dataset was employed to account for this variability. Horizontal and vertical flips of the images were included to make the network segmentation invariant to image orientation. Random zoom up to 20%, rotation up to 10%, and shear up to 5% were used to account for variation in magnification and imaging angles. When zooming out, shearing, or rotating the image, a constant black fill was used to mimic images acquired using physical X-ray filters. The combination of the above data augmentations created a training dataset of over half a million images to improve network generalizability. Data augmentation was not applied to the test set. The augmented UM dataset was used for neural network training, and the test set was used to compare segmentation accuracy.

#### 2. MMM Dataset

The percent change in vessel diameter at the region of stenosis is a key determinant of whether a patient requires an intervention or not; therefore, the accuracy of AngioNet’s segmented vessel diameters was assessed in addition to its overall segmentation accuracy. Although the main result of a QCA report is the overall percent change in vessel diameter, these reports also contain measurements of maximum, minimum, and mean diameter in 10 equal segments of the vessel of interest. These diameter measurements in the MMM QCA Dataset were used to evaluate the discrepancies between QCA and AngioNet.

The data provided by the MMM ICRF Cardiovascular Core Laboratory includes independent and detailed analysis of quantitative angiographic parameters (minimum lesion diameter, percent diameter stenosis, reference vessel diameters, lesion length, maximum percent diameter stenosis, etc.) as per American College of Cardiology/American Heart Association standards, through established QCA software (CAAS-5.10.2, Pie Medical Corp). The study protocol for this data (Computer-Assisted Diagnosis of Coronary Angiography) was approved by the Institutional Ethics Committee of the Madras Medical Mission. This data was obtained using an unfunded Materials Transfer Agreement between UM and MMM. Since the data is completely anonymized and cannot be re-identified, it does not qualify as human subjects research according to OHRP guidelines.

To validate the accuracy of AngioNet’s segmented vessel diameters, a MATLAB script was employed for user specification of the same vessel regions as those in the QCA report. Two regions from the QCA report were sampled in each angiogram. The first was the most proximal region, containing the maximum vessel diameter, and the second was the region of stenosis (given in the QCA report), if present. If no stenosis was reported, the region containing minimum diameter was selected. A skeletonization algorithm ^52^ was used to identify the centerline and radius map of the selected vessel region. Using the output of the skeletonization algorithm, the script reported the maximum and minimum diameters at the selected regions and compared them against the diameters in the QCA report. Maximum and minimum vessel diameter were chosen rather than the diameters on either side of stenosis since the purpose of using the QCA reports was to systematically assess overall vessel diameter accuracy, not the percent diameter reduction. A diagram of the comparison between QCA and AngioNet diameters is shown in Figure 6.

**Figure 6.**
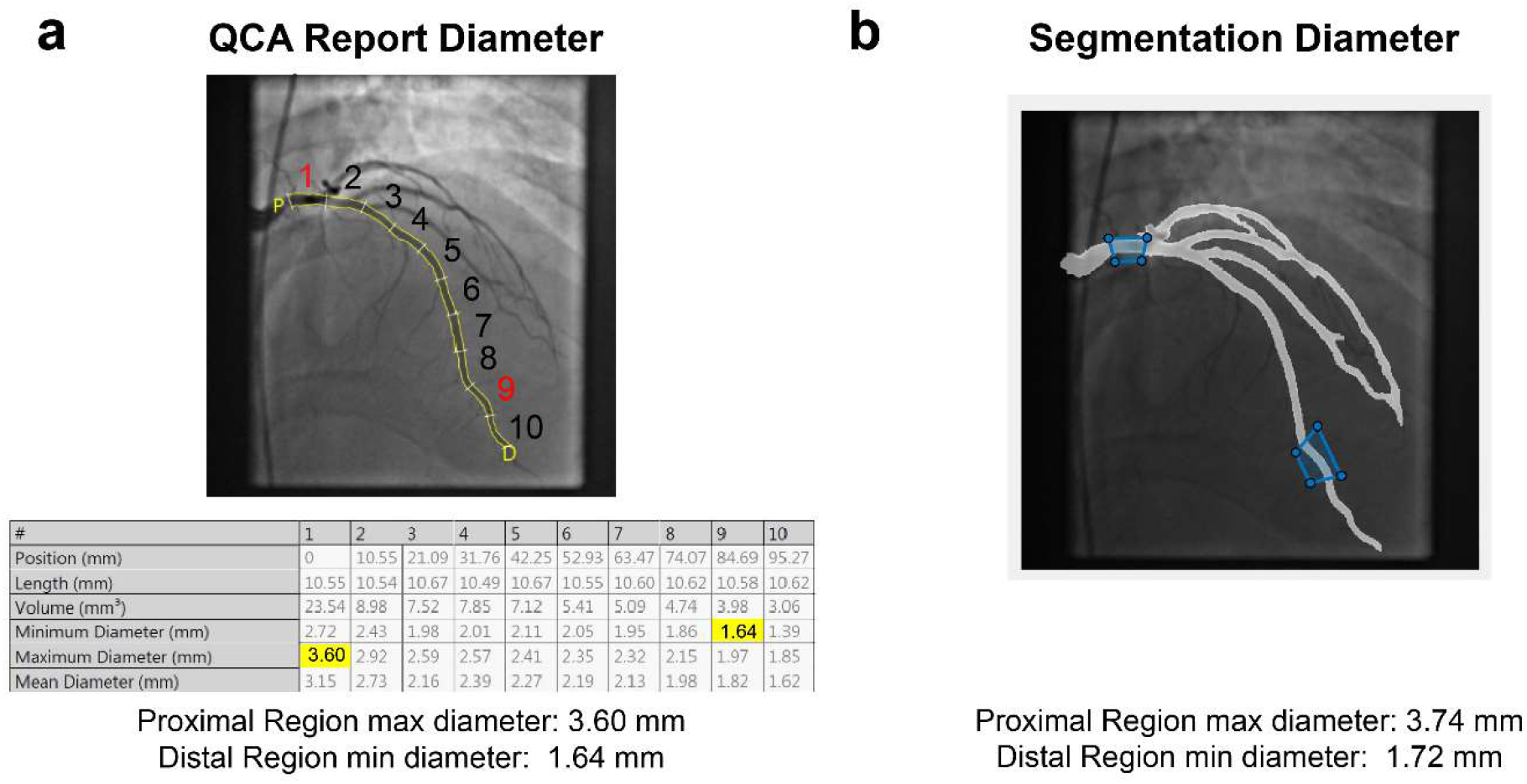
a) Annotated QCA report, along with the corresponding diameters in the report table. Highlighted values correspond to maximum (proximal) and minimum (distal) diameters (segments 1 and 9, respectively). b) Schematic of how a MATLAB script was used to delineate regions in the neural network segmentation corresponding to the regions measured in the QCA report, along with the computed proximal and distal diameters.

### CNN Design and Training

#### 1. Design

AngioNet was created by combining Deeplab v3+ and an Angiographic Processing convolutional neural network (APN). A diagram of the network architecture is given in Fig. 3. Each component of AngioNet, the APN and Deeplabv3+, was trained separately before fine-tuning the entire network.

The purpose of the APN was to address some of the challenges specific to angiographic segmentation, namely poor contrast and the lack of clear vessel boundaries. The APN was initially trained to mimic a combination of standard image processing filters instead of initializing with random weights, since it would later be fine-tuned with a pre-trained backbone network. A combination of unsharp mask filters was chosen as these can improve boundary sharpness and local contrast at the edges of the coronary vessels, making the segmentation task easier. Other forms of preprocessing were also considered, including Contrast Limited Adaptive Histogram Equalization (CLAHE) and singular value decomposition (SVD) denoising. CLAHE has previously been used as an image preprocessing step for angiographic segmentation ^25^, but this method only improves contrast without improving boundary sharpness. SVD denoising was explored as this method could be used to remove imaging artifacts such as filters, patient ribs, or the catheter. While successful in removing these artifacts, SVD denoising also reduced the image sharpness and removed portions of the vascular tree in some images. Therefore, AngioNet’s APN was designed with unsharp mask filters in mind due to their clear advantages in image preprocessing. Using unsharp masking as an initialization, the training process was used to learn a new filter that was best suited for angiographic segmentation.

#### 2) Training

The Deeplabv3+ CNN architecture was cloned from the official Tensorflow Deeplab GitHub repository, maintained by Liang-Chieh Chen and co-authors ^42^. The network was initialized with pre-trained weights from the same repository, as recommended by the authors for training on a new dataset. The input to this network were raw angiographic images, and the output was a binary segmentation. Training was conducted using four NVIDIA Tesla K80 GPUs on the American Heart Association Precision Medicine Platform (https://precision.heart.org/), hosted by Amazon Web Services. Hyperparameters such as batch size, learning rate, learning rate optimizer, and regularization were tuned. We observed that training with larger batch size led to better generalization to new data. A batch size of 16 was used as this was the largest batch size we could fit into memory using four GPUs. The Adam optimizer was chosen to adaptively adjust the learning rate, and L2 regularization was used to reduce the chance of over-fitting. The vessel pixels account for 15-19% of the total pixels in any given angiography image. Due to this class imbalance, it was important to encourage classification of vessel pixels over background using weighted cross-entropy loss^53^. The APN was initially trained to mimic the output of several unsharp mask filters applied in series (parameters: radius = 60, amount = 0.2 and radius = 2, amount = 1). This ensured the APN architecture was complex enough to learn the equivalent of multiple filters with sizes up to 121×121 using only 3×3 and 5×5 convolutions. The number of 3×3 versus 5×5 convolutions as well as the network width and depth were adjusted until the APN could reproduce the results of the serial unsharp mask filters. Additionally, the combination of standard filters was hypothesized to be a good initialization before training the network to learn the best possible preprocessing filter. The inputs to the APN were the normalized images from the augmented UM Dataset, whereas the output was a filtered version of the image. The ground truth images were generated by applying several unsharp mask filters with various parameters to each normalized clinical image. The APN was composed of several 3×3 and 5×5 convolutional layers (Figure 7) and was trained to mimic the unsharp mask filters by minimizing the Mean Squared Error (MSE) loss between the prediction and ground truth images. The APN design and training were carried out using TensorFlow 2.0, integrated with Keras ^54,55^.

**Figure 7.**
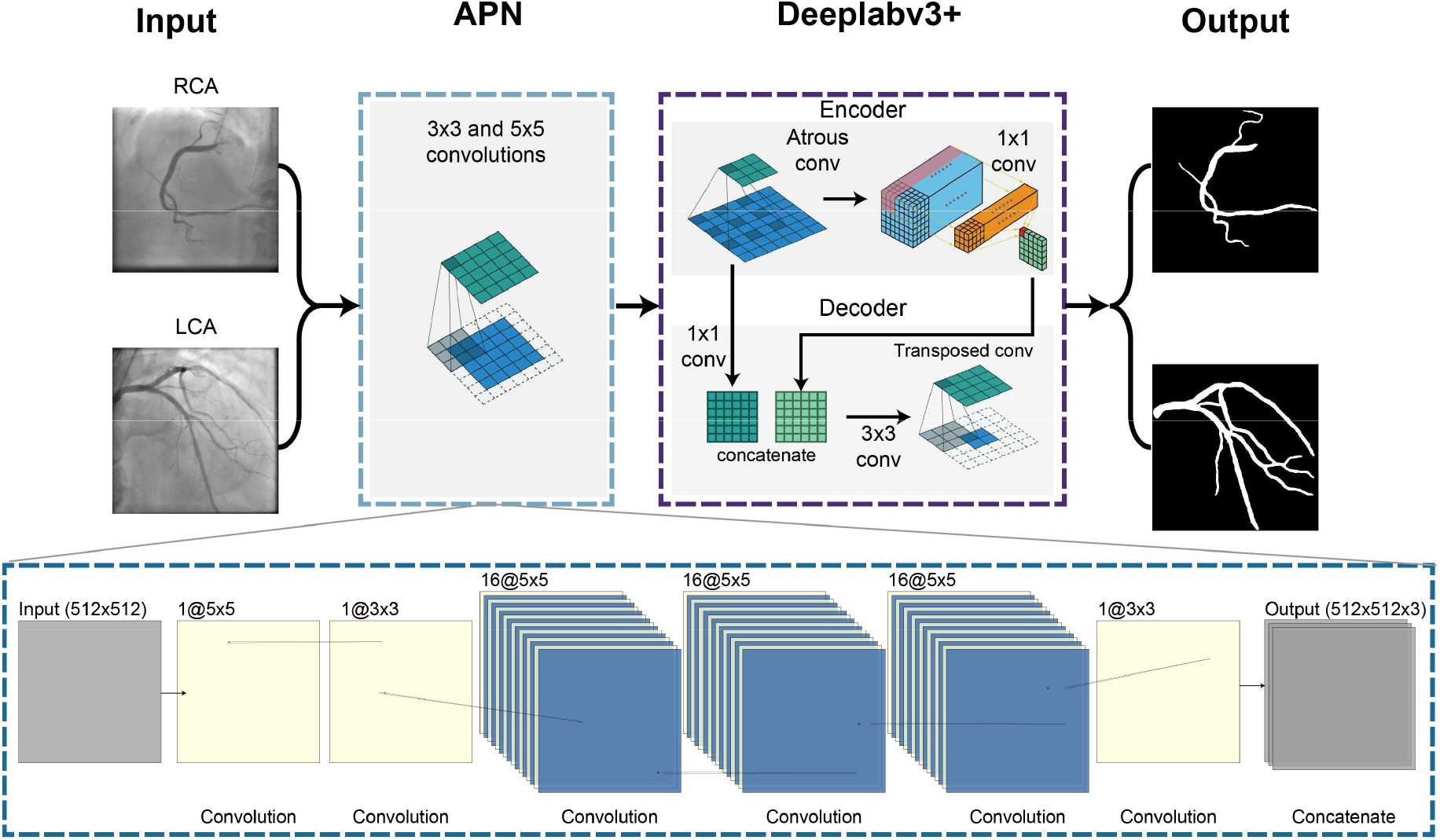
AngioNet Architecture Diagram. AngioNet is composed of an Angiographic Processing Network (APN) in tandem with Deeplabv3+. The APN is designed to improve local contrast and vessel boundary sharpness.

Once the APN and Deeplabv3+ networks were individually trained, the two CNNs were combined to form AngioNet using the Keras functional Model API ^56^ (Figure 7). The resulting network was trained with a low learning rate to fine-tune the combined model. Since neither the APN nor Deeplabv3+ were frozen during fine-tuning, both were able to adjust their weights to better complement each other: the APN learned a better filter than its unsharp mask initialization, and Deeplabv3+ learned the weights that could most accurately segment the vessel from the output of the APN. Batch size, regularization, learning rate, and learning rate optimizer parameters were once again tuned. The same process of pre-training, combining models, and fine-tuning was carried out with the APN and U-Net to determine how much the backbone network contributes to segmentation performance. U-Net was not initialized with pre-trained weights as our dataset was adequately large to train this network from random initialization.

During all phases of training, batch normalization layers were frozen at their pre-trained values as we did not have a large enough dataset to retrain these layers. Furthermore, all hyperparameter optimization was performed on the 5-fold cross validation holdout set and accuracy was measured on the test set.

## Data Availability

Since the datasets used in this work contain patient data, these cannot be made generally available to the public due to privacy concerns. The code for the AngioNet architecture and examples of synthetic angiograms are available at https://github.com/kritiyer/AngioNet. This code is licensed under a Polyform Noncommercial license.

## Acknowledgment

Funding for this work was provided by a grant from the American Heart Association [19AIML34910010]. KI was funded by the National Science Foundation Graduate Research Fellowship Program [DGE1841052] and Rackham Merit Fellowship, and CAF was supported by the Edward B. Diethrich Professorship. CJA was funded by a King’s Prize Research Fellowship via the Wellcome Trust Institutional Strategic Support Fund grant to King’s College London [204823/Z/16/Z].

The authors would like to thank the American Heart Association Precision Medicine Platform (https://precision.heart.org/), which was used for data analysis. Neural network prototyping and initial testing were carried out on a Titan XP GPU granted to our lab by the NVIDIA GPU grant program.

